# COVID-19 Outbreak, Social Response, and Early Economic Effects: A Global VAR Analysis of Cross-Country Interdependencies

**DOI:** 10.1101/2020.05.07.20094748

**Authors:** Fabio Milani

**Author notes:** Address*: Fabio Milani, Department of Economics, 3151 Social Science Plaza, University of California, Irvine, CA 92697-5100. *E-mail*. *Homepage*: http://www.socsci.uci.edu/~fmilani.

## Abstract

The COVID-19 pandemic underscored the importance of countries’ interconnections in understanding and reacting to the spread of the virus.

This paper uses a global model with a sample of 41 countries to study the interdependencies between COVID-19 health shocks, populations’ risk perceptions about the disease, and their social distancing responses; it also provides some early evidence about potential economic effects.

Social networks are a central component in understanding the international transmission.

We exploit a dataset on existing social connections across country borders, made available by Facebook, and show that social networks help explain not only the spread of the disease, but also cross-country spillovers in risk perceptions and in social behavior. Social distancing responses across countries are measured based on aggregated mobility tracking indicators, obtained from Google Mobility Reports.

We estimate a Global VAR (GVAR) model, which allows for endogeneity of each health, social, and economic, domestic variable, and for a dependence of domestic variables on country-specific foreign aggregates, which depend in turn on the matrix of social connections.

Our empirical results highlight the importance of cross-country interdependencies and social networks. Risk perceptions and social responses are affected by experiences abroad, with Italy and the U.S. playing large roles in our sample. The social distancing responses to domestic health shocks are heterogeneous across countries, but they share some similarities: they adjust only gradually and with delay, hence displaying adaptive behavior.

Early indicators are suggestive of unemployment consequences that vary widely across countries, depending on their labor market characteristics. Unemployment is particularly responsive to health shocks in the U.S. and Spain, while the fluctuations are attenuated almost everywhere else.

## 1. Introduction

After being identified in December in Wuhan, China, the novel coronavirus (SARS-CoV-2) initially spread to the Hubei region and later to mainland China. Although the rest of the world soon learned about the first publicly known cases, several countries didn’t perceive an immediate risk for their populations. Starting already in January, the epidemic spread outside China, first in Thailand, South Korea, Japan, and in the United States, and in many cases it was connected to recent travelers to the country. In Europe, Italy had the first official community-based case on February 20, and, very quickly, clusters of cases developed in the Lombardy region. It was later discovered that the virus had been circulating in Lombardy since at least early January (Cereda et al., 2020), and, possibly, since December. From mid-March, the vast majority of countries in the world had multiple cases, with the centers of the outbreak moving first to Europe and, later, to the United States.

Most countries responded by requiring their populations to adhere to some form of social distancing, to reduce the rate infection and lessen the strain on healthcare providers. The responses, however, have been widely heterogeneous. Italy reacted with a few-days delay after the outbreak and then implemented restrictive stay-at-home policies. A minority of countries initially experimented with laxer restrictions, either based on a misguided attempt to have their populations achieve herd immunity on their own (the U.K., which soon moved away from the policy), or because of an unwritten ‘social contract’ with citizens, rather than enforcement from policymakers (Sweden). Others acted quickly and decisively to attempt eradicating the disease before it became widespread (New Zealand).

The spread of Coronavirus has highlighted the importance of interdependencies across different regions. Depending on business links and other existing relationships, the virus rapidly moved across borders. Perceptions about the crisis and social behavior responded generally with lags, but they were also likely affected by the observed experiences abroad. Countries had the opportunity of learning from others about social adjustments that were more or less effective in containing the disease.

The main objective of this work is to study these global interrelationships in the response to COVID-19 shocks. In particular, this paper exploits information about social networks across countries to study interdependencies in the number of disease cases, in the perceptions of their citizens about coronavirus’ risk, and in their social responses. We also provide some preliminary evidence on the early economic effects of the pandemic, by looking at a potential leading indicator of unemployment.

We include in our sample 41 countries and we use a variety of data sources. To measure the extent of pairwise country social connections, we use data obtained from Facebook, which measure the total number of friendships across pairs of countries, as a fraction of the total number of combined users in the two countries. This Social Connectedness indicator allows us to have a measure that can account for different types of relationships: regular friendships, business links, family ties, relations based on older and more recent patterns of immigration, and tourism flows. Social networks can help explain the transmission of COVID-19 cases across borders, and they are likely to represent a superior measure compared with the use of geographic distance alone.^1^ Other contemporaneous papers make a similar observation (e.g., Kuchler, Russel, and Stroebel, 2020). At the same time, social networks not only can potentially explain patters of disease contagion, but also spillovers in ideas and behavior. Controlling for the country-specific dynamics of COVID-19 cases, people’s risk perceptions may respond differently and also be affected by the experience and perceptions of individuals in their networks of social connections, including those residing abroad. The same is true for responses in terms of social distancing: individuals who had large connections to countries where the virus outbreak and the social distancing responses were already happening, may have learned from their early experiences, taken the epidemic more seriously, and responded similarly.

To measure the social distancing response, we exploit a novel dataset made available by Google, through its country-specific Social Mobility reports. The Google data measure changes in mobility calculated using anonymized, aggregated, GPS location tracking from mobile devices, for users who have the location history tracking allowed. The data are computed for each day during the pandemic and compared with corresponding values in the same day of the week for a pre-pandemic sample. Changes in mobility refer to a variety of locations: workplaces, transit stations, entertainment venues, retail stores, parks, and so forth.

We measure coronavirus risk perceptions using information about online searches related to Coronavirus (for example, using the Coronavirus Topic function, we also include searches as ‘Coronavirus symptoms’, ‘Coronavirus treatment’, and so on) through Google Trends. Risk perceptions are influenced by the number of cases in the country, but also likely by events elsewhere. Data from Google Trends have become a popular way to measure people’s attention to certain events or to track realtime developments (related to epidemics, diseases, or the economy, for example) that would be hard to measure otherwise. We also analyze the early economic effects implied by the health shocks and the associated social responses. To have a measure that is connected to unemployment, but available daily, we similarly use Google searches about the Unemployment Topic (again, including all searches related to unemployment, such as ‘unemployment benefits’, ‘unemployment insurance’, ‘how to apply for unemployment’, ‘losing my job’, and so forth). Askitas and Zimmermann (2009) and Choi and Varian (2009), among several others, show that unemployment searches provide a useful early real-time indicator for actual unemployment releases, which are, instead, available only at monthly frequency and with a publication lag.

We estimate a Global VAR model to study the transmission of pandemic health shocks both domestically and globally.^2^ In our global framework, for each country, COVID-19 cases can affect risk perceptions, which can trigger the social distancing response. As a result of social distancing, unemployment may increase. The model allows us to treat all these variables as endogenous: this is necessary since social distancing, for example, is likely implemented in response to rising numbers of COVID-19 cases, but it also has itself an impact on the future number of cases. Moreover, domestic variables in the model are also allowed to respond to foreign aggregates. The foreign variables enter each domestic model with weights that depend on the matrix of social connections: the relevant foreign aggregate for each country is different, since the patterns of connections are unique to the country. In the GVAR literature, the domestic models can be estimated separately as conditional VARs. All endogenous variables can then be stacked together to form a large-scale Global VAR; it is then possible to track the responses of all variables to each shock in each country. Through the use of a connectivity matrix (our Social Connection matrix), the Global VAR model offers a relatively simple and parsimonious way to deal with potentially complex interactions across different variables and countries.

### Main results

Our estimates highlight the importance of interdependencies and social networks in the transmission of coronavirus cases, in the increase of risk perceptions, and in social distancing behavior. Domestic variables, for the vast majority of countries, are significantly affected by foreign aggregates, constructed with weights based on the strength of social connections across countries.

Given the role played by Italy and the U.S. as centers of the outbreak in different phases of the epidemic, we study how variables in the rest of the world respond to coronavirus shocks originating in these countries. We document strong and significant responses of risk perceptions and social distancing to the Italy COVID shock almost everywhere in the world. Countries also respond to the subsequent U.S. shock, although with a smaller magnitude.

The countries’ responses to foreign and to their own domestic coronavirus shocks are heterogeneous. We can, however, reveal some common patterns. The countries that respond with social distancing do so with a delayed and sluggish adjustment. They seem to learn from the experience of other countries, but they display an adaptive behavior: they don’t adjust their habits instantly, but they gradually reduce their social mobility reaching a negative peak after less than a week. On the opposite direction of causality, changes in social distancing lead to a decline in the growth rate of COVID-19 cases.

The implications of the pandemic for unemployment also significantly vary by country. Labor markets in the U.S. and Spain are the most negatively affected, with large expected increases in the unemployment rate. But large spikes in unemployment are not a given, as most other countries seem to experience much more contained fluctuations. The results suggest that different institutional features can partly insulate the corresponding populations from the worse effects of large exogenous shocks.

### Related Literatures

Due to the historical importance of the COVID-19 pandemic, research related to the disease and its effects has been growing swiftly. Many papers use the leading model in epidemiology, the SIR (or, alternatively, the extended SEIR) model based on Kermack and McKendrick (1927), to simulate the evolution of the disease (e.g., Ferguson et al., 2020). In economics, a number of recent papers have adopted a similar framework and developed the theory further by adding relevant trade-offs between health and economic costs (e.g., Eichenbaum, Rebelo, and Trabandt, 2020, Alvarez, Argente, and Lippi, 2020, Jones, Philippon, and Venkateswaran, 2020). This paper, instead, takes a different route by providing empirical evidence related to the social response to the outbreak, and using an alternative framework. In contrast to studies using the SIR model, we do not aim to predict the evolution of the number of infected individuals in a population; our focus lies more on explaining the social responses to the original health shocks around the world.

Other recent works investigate the determinants of different approaches to social distancing. Gupta et al. (2020) find that social distancing responses do not necessarily correspond to policies mandated by State and local governments. Painter and Qiu (2020) and Adolph et al. find that political beliefs affect compliance with social distancing indications in the U.S. Andersen (2020) finds evidence of substantial voluntary social distancing and he also shows that it is affected by partisanship and media exposure.

Our paper provides a contribution to the literature on GVAR models (see Chudik and Pesaran, 2016, for a survey). Most papers in the literature consider macroeconomic applications and study the global spillovers of policy and other shocks (e.g., Pesaran et al., 2004, Chudik and Fratzscher, 2011, Dees et al., 2007). Others have studied interdependencies in housing markets (Holly, Pesaran, and Yamagata, 2011), firm-level returns (Smith and Yamagata, 2011), and a variety of other applications (Di Mauro and Pesaran, 2013). The effect of foreign variables is usually made to depend on trade balances across countries. Our framework, instead, introduces a different connectivity matrix, based on social networks, which can be promising for a different set of applications. Therefore, our paper is also connected to recent papers that propose the use of Facebook connections to measure social networks across locations (Bailey et al., 2018).

Finally, we measure risk perceptions and fears of unemployment using Google Trends data. This approach has become more and more popular, and is now exploited in different fields, to measure people’s attention (Da et a., 2011), in forecasting and nowcasting economic variables (see the various examples discussed in Choi and Varian, 2012), and to track the spread of diseases (e.g., Ginsberg et al. 2009, Brownstein et al., 2009) in the absence of easily observable private information. In our paper, search data help us measure both people’s perceptions about coronavirus and to have a daily indicator of unemployment.

## 2 COVID-19 and Social Response Data

The paper exploits a variety of newly available datasets to study the interrelationship between health shocks originating from the Covid-19 pandemic, people’s real-time perceptions about coronavirus risk, the extent of their social distancing response, and unemployment. We investigate the connections among these variables both within countries, and across borders, by studying contagion and spillovers internationally.

The data are collected on a sample of 41 countries. Those include current OECD member countries, candidate countries that applied for membership, and the countries that the OECD defines as key partners (Brazil, India, Indonesia, South Africa).^3^

Data on novel COVID-19 cases each day for each country are made available by Johns Hopkins University’s Center for Systems Science and Engineering (CSSE). In our estimations, we use either the growth rate or, for robustness, the number of daily cases.

The epidemiology literature stresses the importance of social distancing to contain the spread of the virus, by reducing the basic reproduction number *R*_0_ (the expected number of secondary infections produced by a single infection in a population where everybody is susceptible) and flattening the curve of infected individuals. The response has been different across countries, either in terms of policies, enforcement, or voluntary reductions in mobility. Therefore, it’s important to have accurate data on actual social distancing by different populations to track the implied health and economic effects. To this scope, we use daily time series indicators on Social mobility made available by Google. The indicators are obtained using aggregated, anonymized, data from GPS tracking of mobile devices, for users who opted in to ‘Google Location History’.

The data measure the change in the number of visits and length of stay at different places compared to a baseline. For each day of the week, mobility numbers are compared to an historical baseline value, given by the median value for the corresponding day of the week, calculated during the five-week period between January 3 and February 6, 2020. The data are reported for five place categories: Grocery and pharmacies, parks and beaches, transit stations, retail and recreation, and residential.

In addition to the official number of COVID-19 cases, which may be an imperfect measure of the pervasiveness of the virus in the population, we also measure the population’s risk perception about coronavirus. The risk perception is measured using daily data on web searches from Google Trends. We use the search results for the whole ‘Topic’ category, therefore, our indicator also include all related search terms as ‘Coronavirus symptoms’, ‘Coronavirus treatment’, ‘Coronavirus vs. flu’, and so forth.

Finally, we similarly use an indicator of unemployment to measure the initial economic effects of the outbreak. Given that actual unemployment data are typically monthly and their release lagged by more than a month, we also exploit Google Trends data about unemployment as a variable that can be used to have early, real-time, indications of the official variable. Askitas and Zimmermann (2009) and Choi and Varian (2009) show that unemployment searches are can help predict initial unemployment claims and the unemployment rate. Our unemployment variable can be interpreted as a real-time signal for unemployment, or, alternatively, as a measure of people’s perceptions, attention, or fears, about unemployment over the time period that we study.^5^

Finally, we measure international social connections using Facebook’s Social Connectedness data. The index uses active Facebook users and their friendship networks to measure the intensity of connectedness between each pair of locations. The measure of Social Connectedness between two locations *i* and *j* is given by:

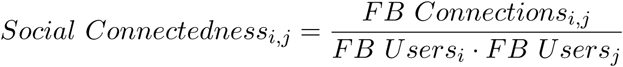

where *FB Connectionsi j* denotes the number of friendship connections between region *i* and *j*, and *FBUsers_i_, FBUsers_j_* denote the number of Facebook users in *i* and *j*. The Social Connectedness index, therefore, measures the relative probability of a Facebook connection between any individual in location *i* and any individual in location *j*. The data used in this paper refer to the measure calculated for March 2020.

Bailey et al. (2018) proposed the measure to study the effects of social networks across U.S. counties. Other current papers are uncovering the link between social networks and the diffusion of Covid-19 (e.g., Kuchler et al., 2020). The measure can be preferred to alternatives based simply on inverse geographic distance, since it can provide a more accurate account of business relations, tourism patterns, and family or friendship ties, across different areas. We argue here that the strength of social connections can also affect information about the outbreak and social distancing responses.

Figures 1 and 2 show the likelihood of social connections across countries, with Italy and the U.S. chosen as examples (and, therefore, shown in red in their corresponding figure).

**Figure 1:**
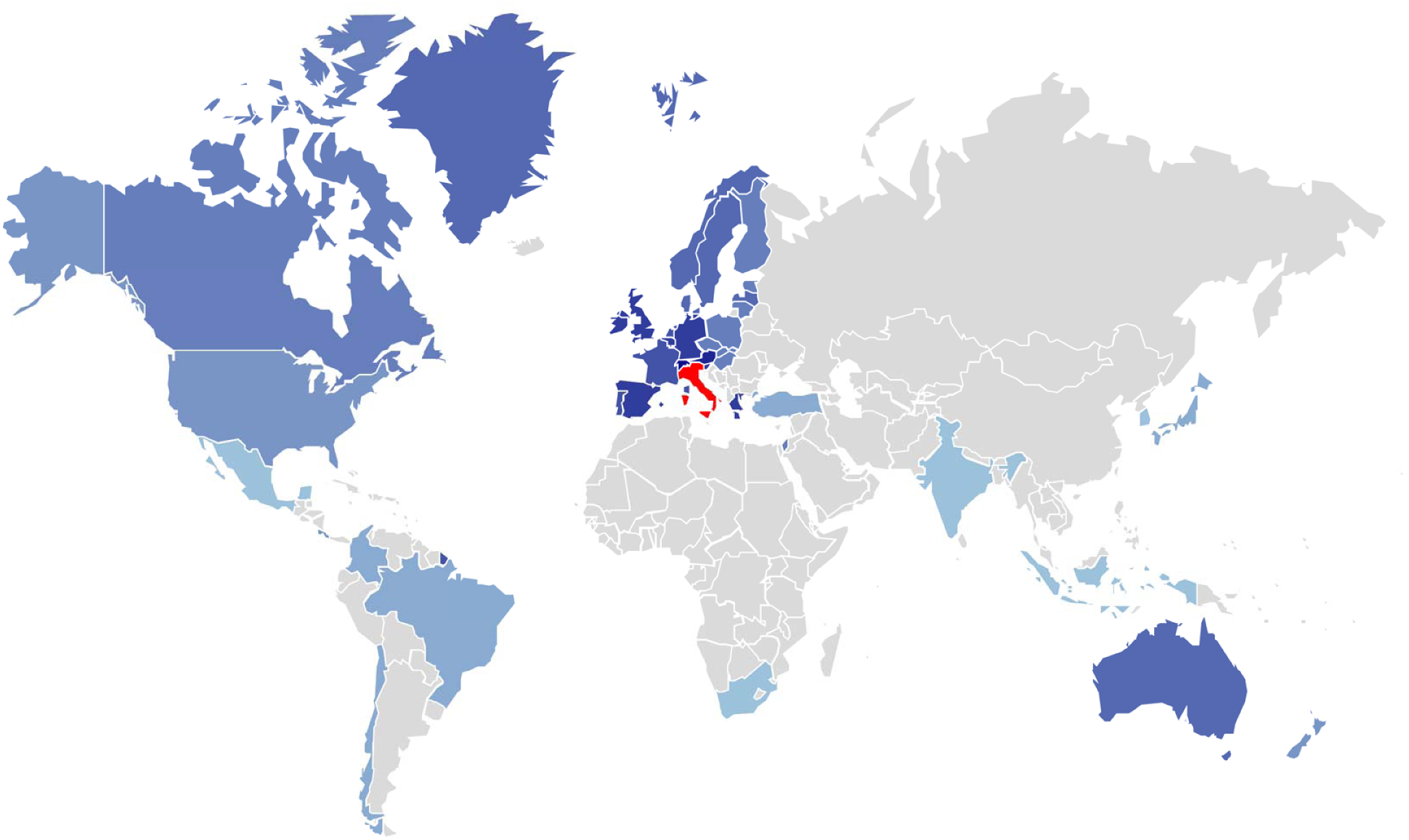
Social Connections between Italy and the rest of countries in the sample. *Note*: the reference country (Italy) is shown in red; social connections are measured with different tonalities of blue, with darker tones denoting stronger connections; countries that are not considered in our estimation are left in grey.

**Figure 2:**
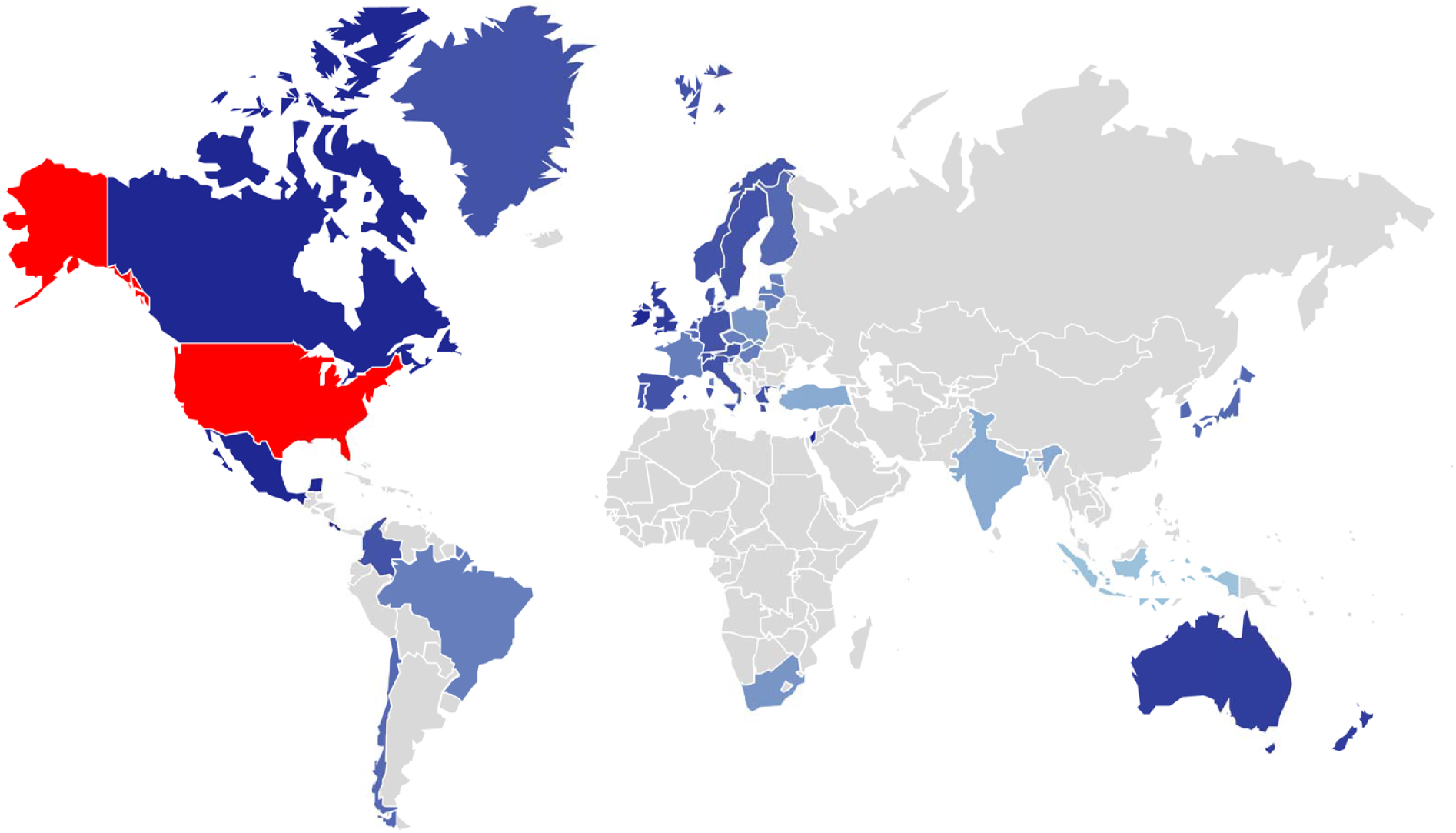
Social Connections between the United States and the rest of countries in the sample. *Note*: the reference country (U.S.) is shown in red; social connections are measured with different tonalities of blue, with darker tones denoting stronger connections; countries that are not considered in our estimation are left in grey.

For Italy, the strongest social connections are with Switzerland and Slovenia, followed by Austria, Germany, Spain, Belgium, and the U.K. Distance is clearly a determinant of social networks, but not the only determinant. Social connections are stronger between Italy and Australia, Italy and the U.S., and Italy and Canada, than between Italy and Turkey, although the latter is geographically much closer.

For the United States, as expected, the most socially connected countries are Mexico and Canada, followed, at lower levels, by Ireland and Israel. The U.S. have strong connections with Australia and New Zealand, which would be downplayed based on a pure measure of distance.

Figure 3 shows, instead, the social distancing response across a sample of major countries in our sample (for easiness of exposition, we show the experiences of 15 out of 41 countries in the figure). Mobility declined by 60% or more in Italy, France, Spain, and New Zealand. While in some countries, the adjustment was abrupt (e.g., New Zealand, France, Spain), it was slower and more gradual in others, such as the U.K. (where the response starts a few days later) and the U.S.; their overall declines in mobility were also more modest. Sweden is an outlier in Europe, as it maintained only small fluctuations of mobility around the historical mean. Japan and Korea observed their first cases earlier, therefore their social distancing responses in our period appear more limited.

**Figure 3:**
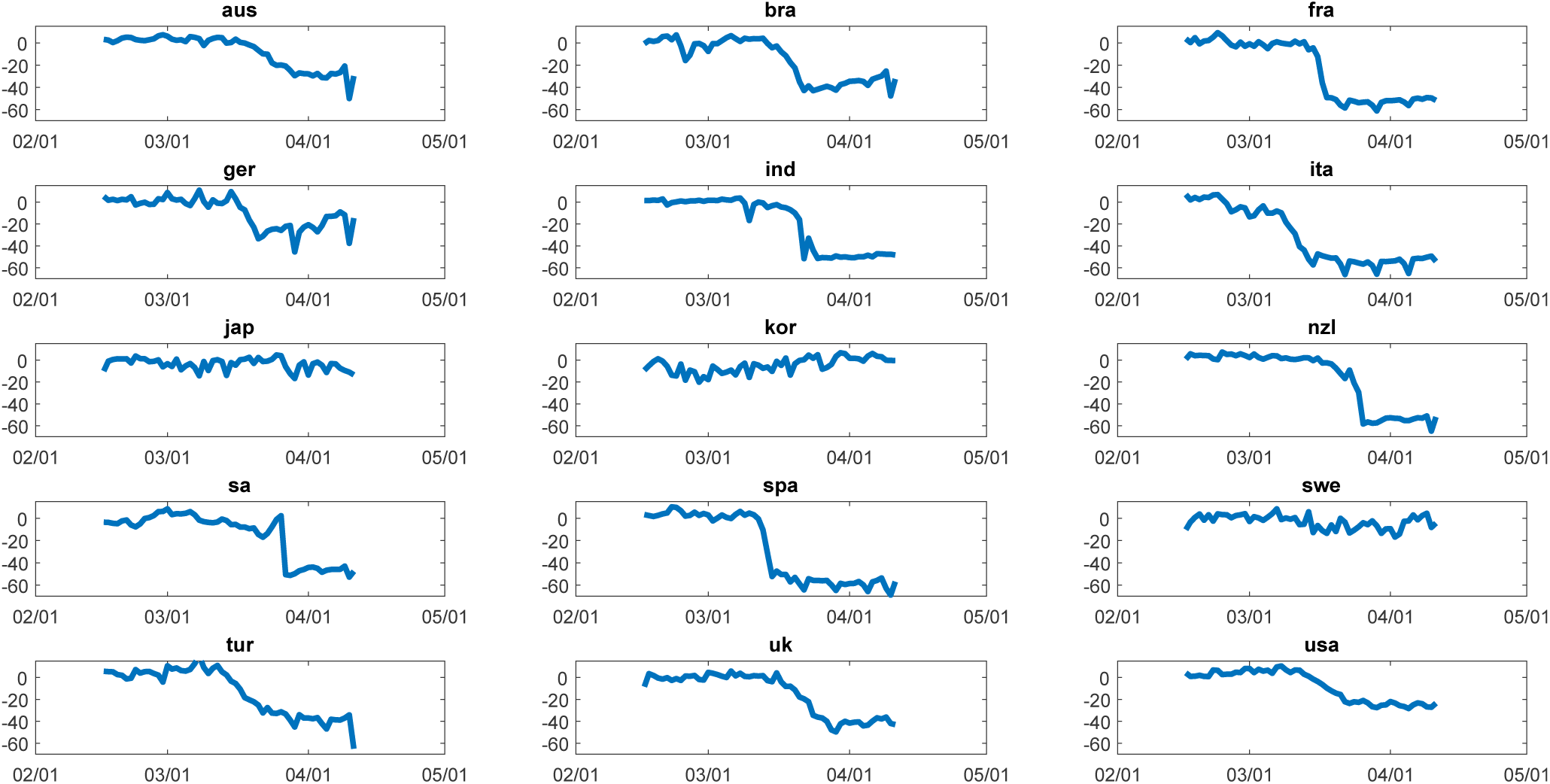
Decline in Social Mobility across a selection of countries (Google mobility data).

## 3 A Global VAR Model

To model global interdependencies in the spread of the disease and countries’ responses, we follow the GVAR approach proposed in Pesaran et al. (2004) and surveyed in Chudik and Pesaran (2016).

Assume that there are *N* units, representing countries in our case, and for each unit, the dynamics is captured by *k_i_* domestic variables. For each country *i*, the *k_i_ ×* 1 vector *x_i_,_t_* of endogenous variables is modeled as:

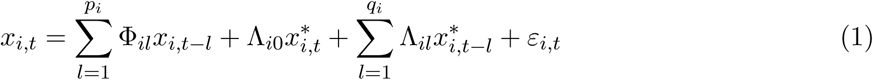

for *i* = 1,2,…,*N* and where Φ*_il_*, 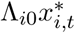, and Λ*_il_* denote matrices of coefficients of size *k_i_* × *k_i_* and 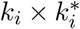, where 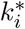 denotes the number of ‘foreign’ variables, and *ε_i_,_t_* is a *k_i_* × 1 vector of error terms. In the empirical analysis, we select the optimal number of lags *p_i_* and *q_i_* for each country using Schwartz’s Bayesian Information Criterion (BIC).

For each country *i*, therefore, domestic variables are a function of their *p_i_* lagged values, possibly of the contemporaneous values and *q_i_* lagged values of foreign, or global, variables. The foreign variables are 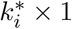 cross-section averages of foreign variables and they are country-specific:

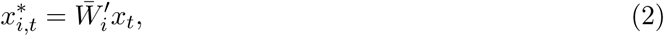

for *i* = 1,…,*N*. The matrix 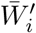 contains country-specific weights, with diagonal elements *w_ii_* = 0. In our approach, we use the extent of social connections across country borders from the Facebook Social Connectedness index dataset to measure the weights.

GVAR models assume that the variables 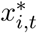 are weakly exogenous. This corresponds to the popular assumption in open-economy macro models that the domestic country is treated as ’small’ in relation to the world economy, i.e, it doesn’t affect global variables. This assumption can be easily tested for all the variables. For cases in which a domestic variable has an unduly large effect on global variables, weak exogeneity will not be invoked there and the foreign variable, instead, will not included in that VAR.

The estimation works in two steps. First, VARX^*^ (that is, VARs with weakly exogenous foreign variables) models can be estimated for each country separately. Second, the estimated country models are stacked to form a large GVAR system, which can be solved simultaneously.

Domestic and foreign variables are stacked in the *k_i_* + *k\** vector 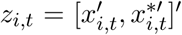. The model in (1) can be rewritten as

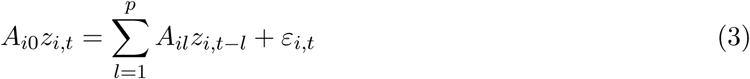

where *A_i_*_0_ = (*I_ki_* − Λ*_i_*_0_), *A_il_* = Φ*_il_*Λ*il*, for *l* = 1,…,*p*, with *p* = *max(p_i_,qi)*. By defining the ‘link’ matrices 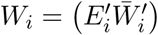, where *E_i_* is a selection matrix that selects *x_i_,_t_* from the vector *x_t_*, we have

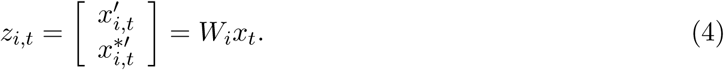

Substituting into (3), and stacking all the unit-specific models, we have

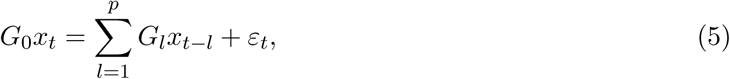

where *G_l_* = *[A*_1_*,_l_W*_1_*,A*_2_*_,l_ W*_2_*,…,A_N,l_W_N_*]′, for *l* = 0,1,…,*p*, and 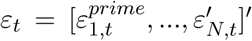. With *G*_0_ invertible, as it is in our case, the GVAR is given by

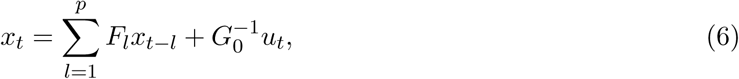

with 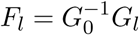.

The GVAR solution can be used to trace the impact of shocks on the variables of interest, both domestically and globally. To find the impulse response to shocks, we adopt the Generalized Impulse Response Function (GIRF) approach, proposed by Koop et al (1996), Pesaran and Shin (1998), and also used in Pesaran et al. (2004). The vector of GIRFs is given by

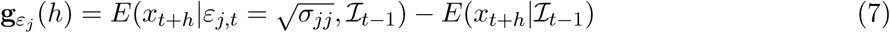

where *j* indexes the different shocks, *h* denotes the horizon for the impulse response function, and where 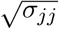 indicates that the magnitude of the shock is set at one standard deviation of the corresponding *ε_j_,_t_*.

The GVAR specification can be seen in relation to a number of econometric alternatives: spatial VARs, panel VARs, and dynamic factor models. Spatial VARs are very strongly connected. They also assume a connectivity matrix, which is usually based on geographic distance. The main difference between the two approaches lies with the structure of correlations: as discussed at length in Elhorst et al. (2018), spatial VARs may be preferred when correlations across units are extremely sparse, for example, when a unit is only affected by few bordering units (“weak”, or local, cross-sectional dependence). The GVAR is meant to capture stronger interrelationships, with dense connectivity matrices, where each country unit is affected, in different ways, by several other countries, or by an aggregate measure (“strong” cross-sectional dependence). Spatial VARs can also be seen as a particularly restricted case of a GVAR model. The approach can similarly be seen as a particular form of panel VAR. The main advantage here is that, through the weight matrix *W_i_*, it allows us to exploit knowledge about social networks and use it to inform the magnitude of cross-country interdependencies. Panel VARs often impose the same coefficients for each unit, shutting down static and dynamic heterogeneity, as well as neglecting cross-country interdependencies. An exception is provided by Canova and Ciccarelli (2009): they introduce a factor structure in the coefficients to solve the curse of dimensionality. Their approach is particularly useful when there is no a priori knowledge that can be exploited about the spillovers. In our case, the extent of social networks can be, instead, exploited to provide some information about the relative strength of interdependencies. Finally, the GVAR has relations with dynamic factor models. As Chudik and Pesaran (2011) show, the GVAR specification approximates a common factor across units, and it extracts common factors using structural knowledge.

The model is particularly suited to account for potentially complex patterns of interdependencies across countries. At the same time, the GVAR specification does so while maintaining simplicity and parsimony. The dimensionality issue is resolved by decomposing a large scale VAR into a number of smaller scale VARs for each unit, which can be estimated separately, conditional on the dynamics of weakly exogenous foreign variables. The interdependencies are not left entirely unrestricted, since it would be unfeasible to estimate all the parameters, but they are given a structure based on knowledge of the data.

We estimate the GVAR model using daily data from February 15 to April 11, 2020. The dates are chosen based on current availability of our Google social mobility data. Each country-specific VAR in expression (1) includes four domestic series in the vector *x_i_,_t_*: the growth rate of Covid-19 cases, the risk perception about Covid-19, the change in social mobility, and the perception about unemployment. The exogeneity assumption is relaxed where it appears unlikely: for Covid cases, we don’t include foreign variables in the model for the US, Italy, and Spain, since they may be endogenous. Those countries, at different times, have accounted for a large share of global cases. We allow the Covid variable for all other countries to be affected by foreign series. We also allow risk perceptions in each country, as well as social distancing outcomes, to be affected even contemporaneously by corresponding variables in different countries. Finally, we assume that domestic unemployment perceptions are affected by foreign unemployment perceptions, but not within the same day. This assumption is not important for the results (which are robust), but it’s motivated by the idea that the unemployment data are driven more by country-specific, than across-the-border, factors. We test the weak-exogeneity for all foreign variables, and they are never rejected in our data.

## 4 Results

### 4.1 Cross-Country Interdependencies

First, to study the magnitude of interdependencies, we show in Table 1 the contemporaneous effects of foreign variables on domestic variables. The Table reports the estimated coefficients, alongside the associated standard errors. Domestic variables are significantly affected by the country-specific foreign aggregates, computed using the matrix of social connections as country-by-country weights. The results indicate that the international spread of COVID-19 cases can be, in part, explained by existing social networks across country borders. Moreover, the contagion is not only physical and related to the spread of the disease, but, even more so, it affects the spread of perceptions and social behavior. Both our measures of risk perceptions in response to the coronavirus threat and the social distancing response are positively influenced by developments in the rest of the world.

**Table 1.** - Contemporaneous effects of foreign aggregates on domestic variables. *Note*: the table report the estimated GVAR coefficients with the associated standard error shown below in parentheses. Significance at the 1% level is denoted by ‘***’, at the 5% level by ‘**’, and at the 10% level by ‘*’.

| Countries | COVID | Risk Perc. | Soc. Dist. | Countries | COVID | Risk Perc. | Soc. Dist. |
| --- | --- | --- | --- | --- | --- | --- | --- |
| Australia | 0.404***<br>(0.094) | 0.726***<br>(0.126) | 1.042***<br>(0.164) | Italy |  | 1.055***<br>(0.389) | 0.643***<br>(0.129) |
| Austria | 0.631***<br>(0.243) | 1.266***<br>(0.117) | 0.902***<br>(0.098) | Japan | 0.081<br>(0.076) | 0.272***<br>(0.091) | 0.239<br>(0.264) |
| Belgium | 0.547***<br>(0.188) | 0.881***<br>(0.095) | 0.781***<br>(0.143) | Korea | -0.524*<br>(0.294) | 0.260***<br>(0.074) | 0.010<br>(0.247) |
| Brazil | 0.999***<br>(0.203) | 0.153<br>(0.172) | 1.068***<br>(0.174) | Latvia | 0.036<br>(0.147) | 0.849***<br>(0.095) | 1.571***<br>(0.191) |
| Canada | 0.326***<br>(0.121) | 1.301***<br>(0.147) | 0.905***<br>(0.179) | Lithuania | -0.008<br>(0.207) | 0.280***<br>(0.076) | 0.989***<br>(0.214) |
| Chile | 1.041***<br>(0.306) | 0.824***<br>(0.153) | 0.809***<br>(0.113) | Luxembourg | 0.032<br>(0.118) | 0.833***<br>(0.163) | 1.084***<br>(0.222) |
| Colombia | 0.512**<br>(0.223) | 0.561**<br>(0.229) | 0.648***<br>(0.200) | Mexico | -0.158<br>(0.287) | 0.463***<br>(0.160) | 0.191<br>(0.190) |
| Costa Rica | 0.316<br>(0.300) | 0.569**<br>(0.223) | 0.755***<br>(0.166) | Netherlands | 1.559***<br>(0.213) | 0.830***<br>(0.089) | 0.696***<br>(0.156) |
| Czech Republic | 0.899***<br>(0.193) | 0.613***<br>(0.112) | 0.679***<br>(0.112) | New Zealand | 0.257<br>(0.209) | 0.562***<br>(0.160) | 0.897***<br>(0.196) |
| Denmark | 0.968***<br>(0.322) | 0.686***<br>(0.130) | 0.845***<br>(0.241) | Norway | 0.761**<br>(0.306) | 0.952***<br>(0.108) | 0.749***<br>(0.292) |
| Estonia | 0.705***<br>(0.263) | 0.555***<br>(0.113) | 1.579***<br>(0.168) | Poland | 0.387<br>(0.252) | 1.522***<br>(0.202) | 0.804***<br>(0.221) |
| Finland | 0.493***<br>(0.101) | 0.725***<br>(0.085) | 0.615***<br>(0.095) | Portugal | 0.283<br>(0.177) | 0.431***<br>(0.133) | 0.277**<br>(0.131) |
| France | 1.046***<br>(0.121) | 0.843***<br>(0.150) | 0.662***<br>(0.095) | Slovakia | 0.131<br>(0.119) | 0.694***<br>(0.143) | 1.268***<br>(0.129) |
| Germany | 1.078***<br>(0.115) | 0.738***<br>(0.196) | 1.640***<br>(0.181) | Slovenia | 0.631***<br>(0.205) | 0.823***<br>(0.101) | 1.152***<br>(0.141) |
| Greece | 1.100***<br>(0.274) | 0.546***<br>(0.108) | 0.361<br>(0.253) | South Africa | -0.104<br>(0.274) | 0.230<br>(0.344) | -0.101<br>(0.361) |
| Hungary | 0.281<br>(0.199) | 0.771***<br>(0.190) | 0.965***<br>(0.162) | Spain |  | 1.560***<br>(0.232) | 1.198***<br>(0.221) |
| India | 1.439***<br>(0.380) | 0.315*<br>(0.167) | 0.511*<br>(0.286) | Sweden | 1.050***<br>(0.265) | 0.782***<br>(0.072) | 0.432**<br>(0.204) |
| Indonesia | 0.864**<br>(0.394) | 0.381**<br>(0.157) | 0.753***<br>(0.116) | Switzerland | 0.739**<br>(0.296) | 0.626***<br>(0.166) | 1.090***<br>(0.239) |
| Ireland | 1.784***<br>(0.337) | 1.860***<br>(0.214) | 0.674***<br>(0.194) | Turkey | 0.277<br>(0.256) | 0.071<br>(0.080) | 0.494*<br>(0.254) |
| Israel | 0.943***<br>(0.247) | 0.246***<br>(0.066) | 0.085<br>(0.381) | U.K. | 0.895***<br>(0.121) | 1.067***<br>(0.186) | 0.587***<br>(0.151) |
|  |  |  |  | U.S.A. |  | 1.622***<br>(0.154) | 0.545***<br>(0.110) |

Only few countries fail to display a statistically significant response to global conditions. Risk perceptions do not rise in response to increasing international distress only in Brazil, South Africa, and Turkey. Most countries also gradually learn from each others’ social distancing responses. Among the few exceptions, Japan and Korea represent a different experience, as they implemented social distancing earlier, but they relaxed already many restrictions in the sample we consider.

### 4.2 Global Transmission of Shocks from Italy and the U.S

We study the global responses to shocks from Italy and the U.S., since these countries played outsized roles in different phases of the pandemic.

Figure 4 and 5 show the impulse response functions for all countries in the sample for the risk perception and social distancing variables to a one-standard-deviation COVID shock originating in Italy. Risk perceptions increase, with some sluggish adjustment, almost everywhere in the world in response to the initial shock from Italy. The peak response usually happens 4-6 days after the shock. The response is more delayed in Brazil, India, and South Africa. Populations in neighboring European countries, as well as in the U.S., Australia, and Canada, significantly update their perceptions. The effect is much smaller in Sweden, Finland, Turkey, Israel, and Lithuania. Again, Japan and Korea don’t seem to significantly respond, as they experienced their outbreaks earlier than the rest of countries. Similarly, most countries respond with declines in social mobility. The social distancing response is already slow and gradual in Italy, with a peak reduction in mobility only 6 days after the shock. Other European countries, as France, Switzerland, and the U.K., don’t seem to adjust at all for the initial 3-5 days, after which they gradually reduce their social mobility as well. The patterns are similar everywhere: while there are strong interdependencies, after a worsening of the situation in one country, others don’t immediately learn and change their behavior. Instead, they behave adaptively: they only slowly and gradually alter their habits in response to the evolving situation.

**Figure 4:**
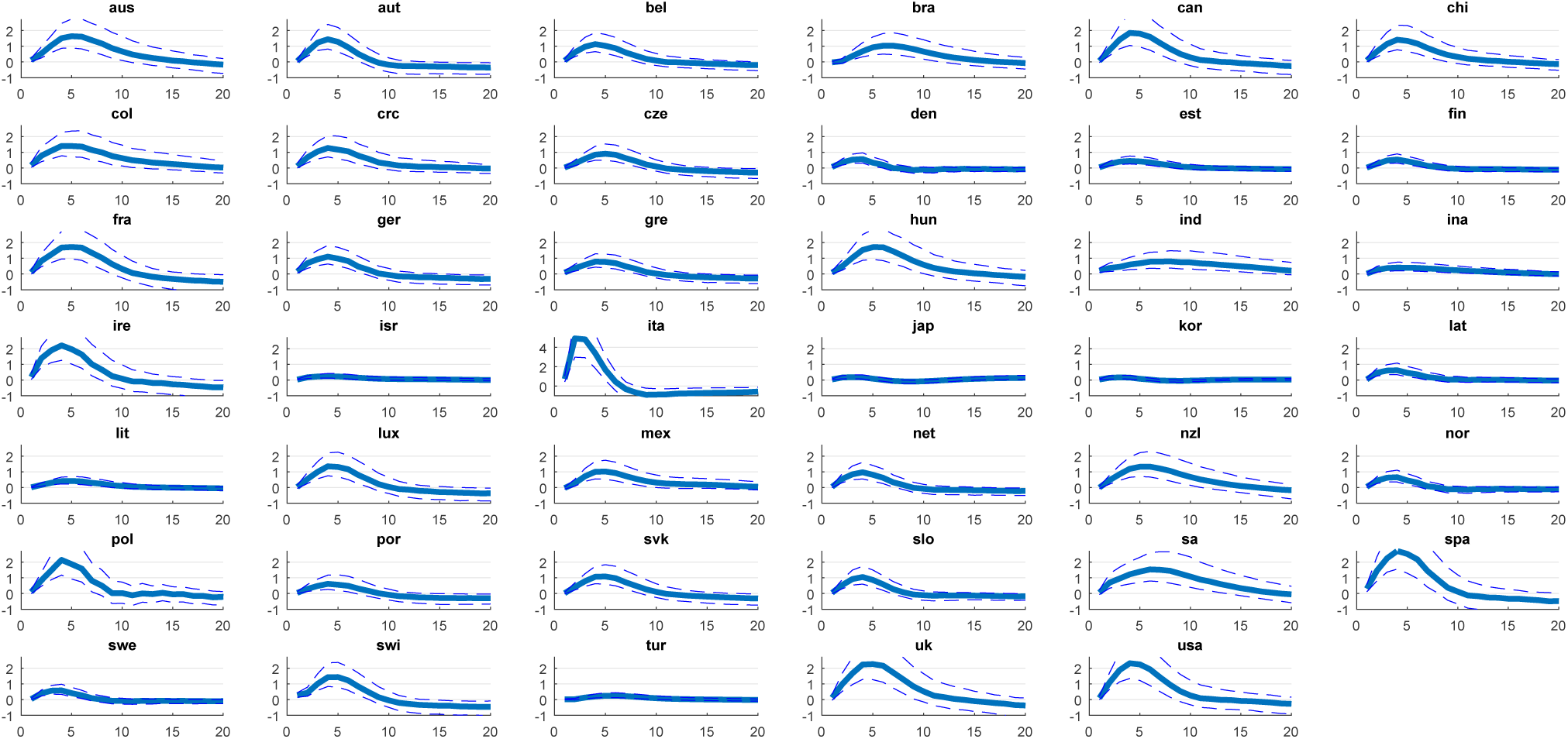
Impulse Responses across countries of Coronavirus Risk Perception to a COVID-19 growth rate shock originating in Italy.

**Figure 5:**
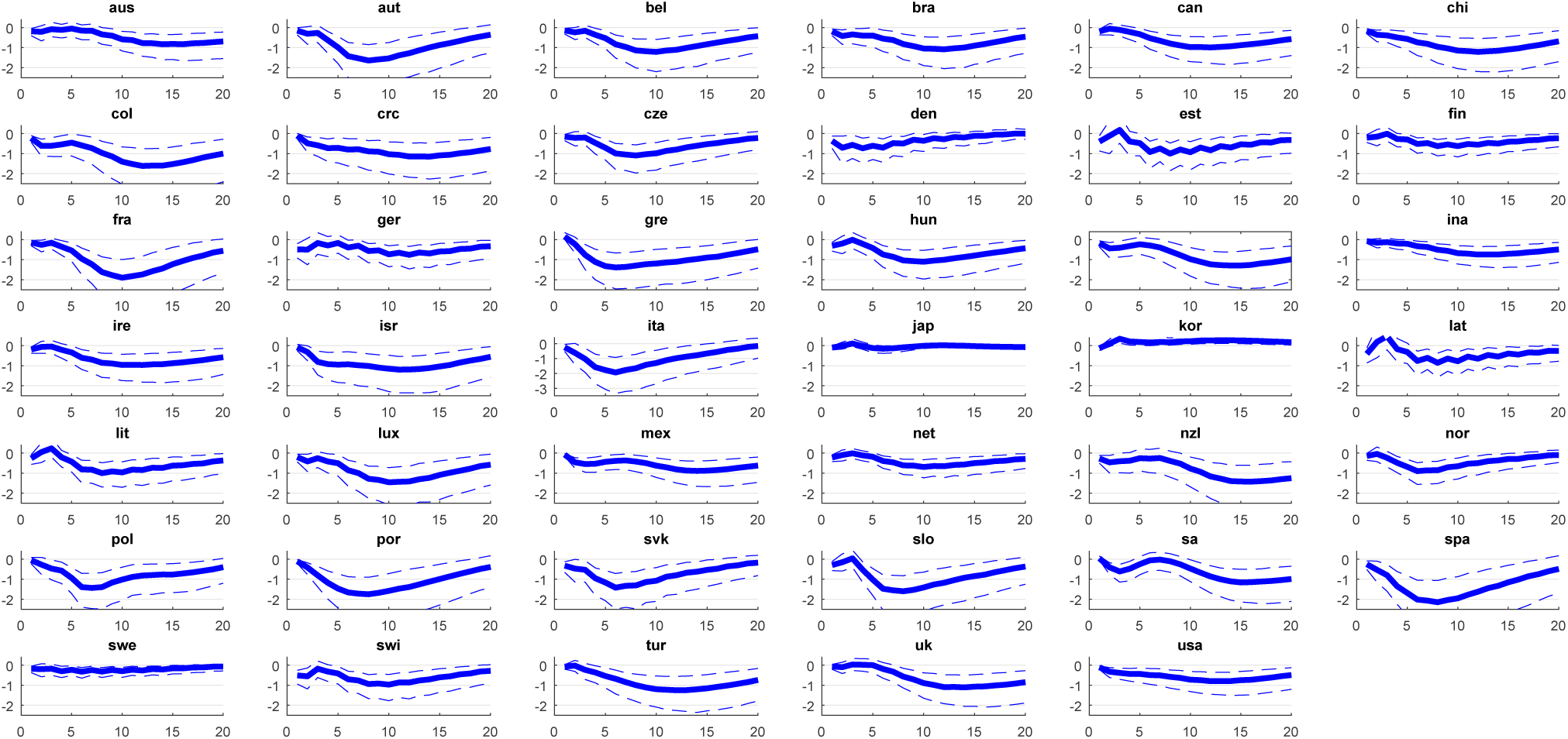
Impulse Responses across countries of Social Mobility to a COVID-19 growth rate shock originating in Italy.

The focal point of the pandemic later moved to the U.S., at least beginning from mid-March. Figure 6 displays the effects on coronavirus risk perceptions in the rest of the world to a U.S. coronavirus risk shock. The spillovers in risk perceptions are again statistically significant, but much smaller in magnitude than those observed in response to the corresponding Italian shock.

**Figure 6:**
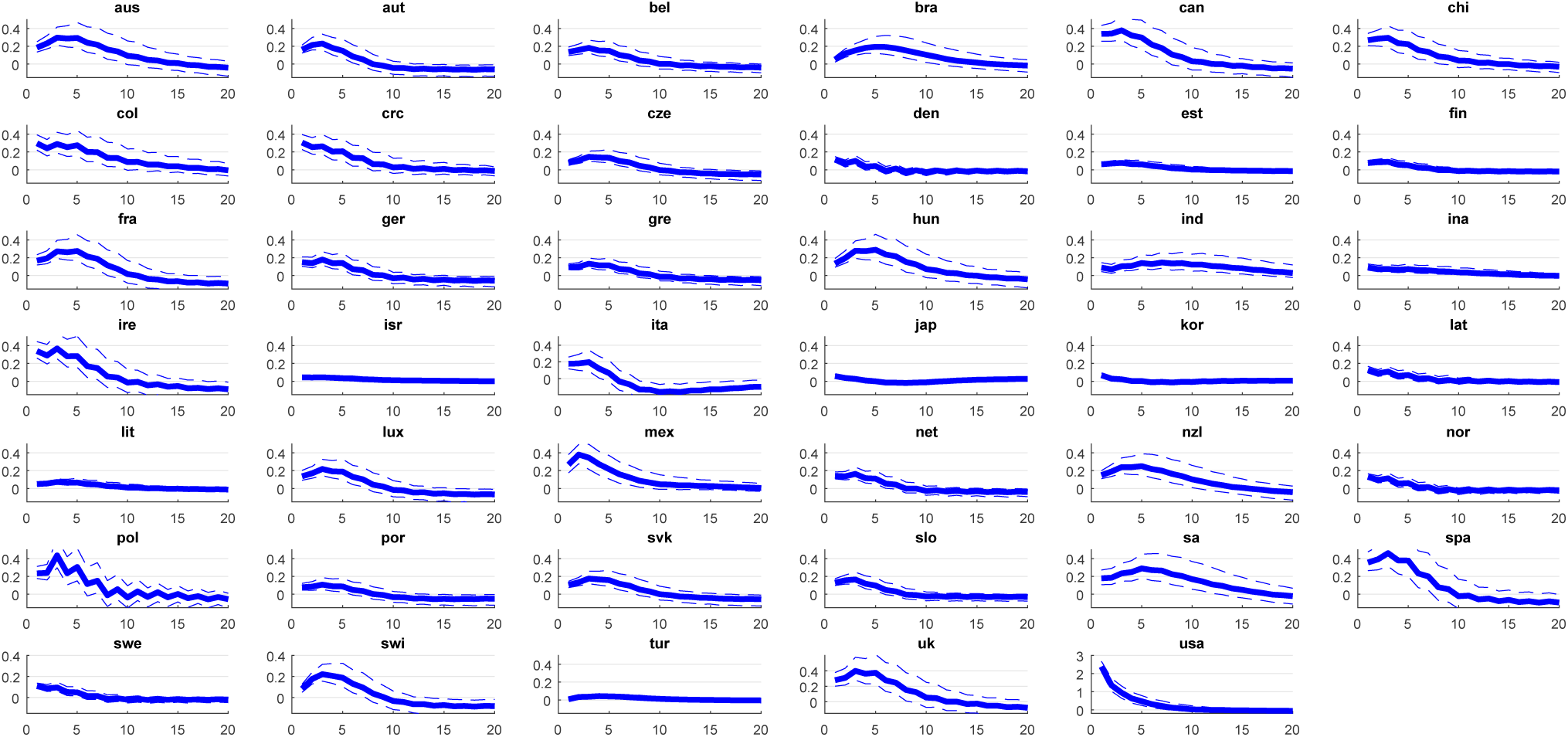
Impulse Responses across countries of Coronavirus Risk Perception to a Coronavirus Risk Perception shock originating from the U.S.

### 4.3 Heterogeneity in Country Responses

The responses to the pandemic have been heterogeneous across countries. Figure 7 overlaps, for a selection of countries, the impulse responses of social distancing and unemployment to the country’s own coronavirus risk shock. We single out the responses for Italy, Spain, the U.K, the U.S., Sweden, and Japan, since they characterize somewhat different approaches to the crisis.

**Figure 7:**
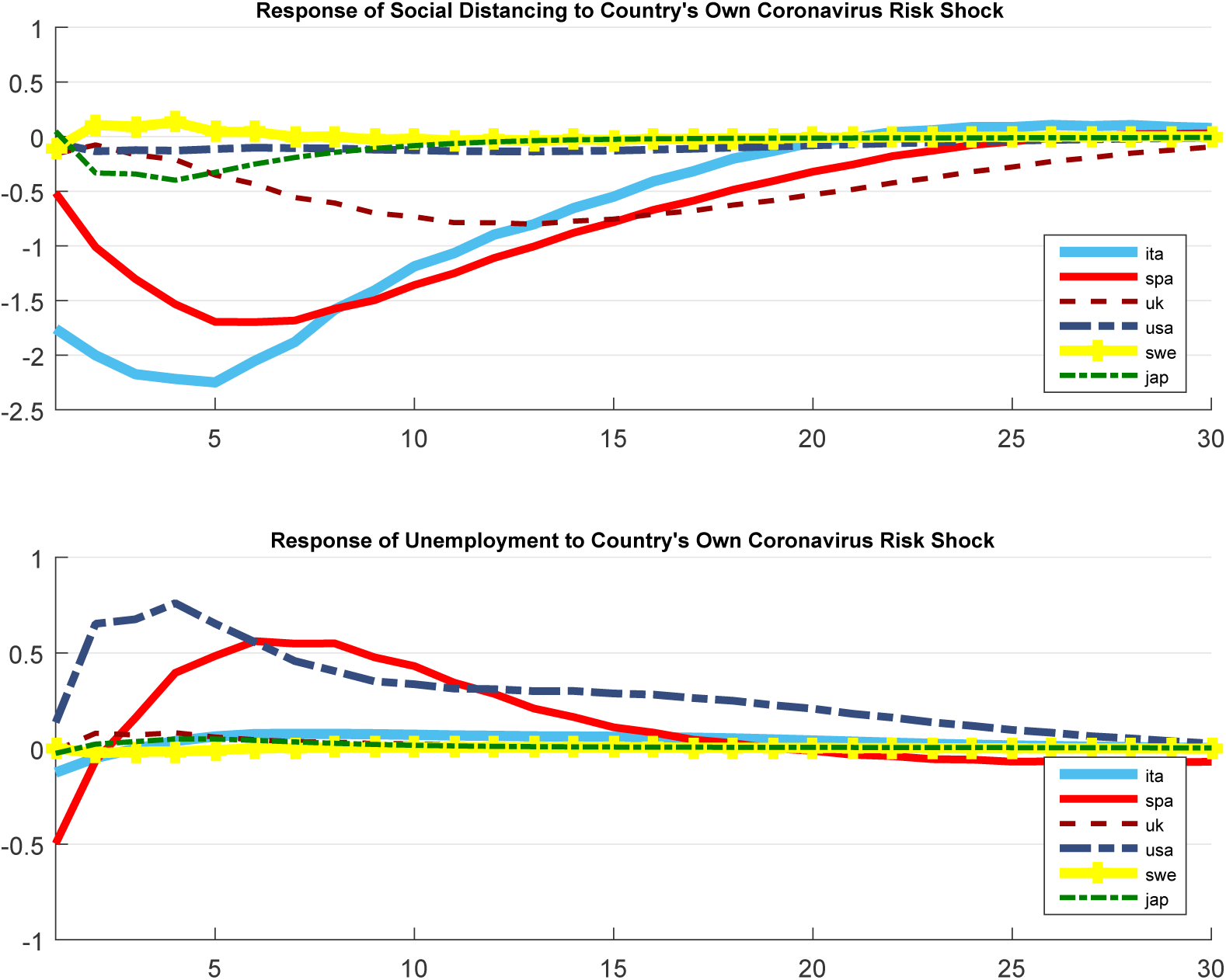
Impulse Response Functions of Coronavirus Risk Perceptions and Unemployment to the Country’s own Coronavirus Risk Shock.

The populations of Italy and Spain strongly decreased their social mobility after the domestic coronavirus shock. The responses reach their maximum effects after roughly five days. Japan display a smaller, more sluggish, response. Although the U.S. and the U.K., if taken by themselves, would also reveal negative and statistically significant social distancing, their responses are many order of magnitudes smaller than the ones observed in Italy and Spain. Finally, it is well documented that Sweden adopted a more permissive approach, by letting its citizens adjust their behavior without the same strict enforcement as in other countries. The response for Sweden, accordingly, doesn’t show any significant decrease in mobility to the country-specific risk shock.

Turning to the early estimates about potential economic effects, we show the response of our real-time unemployment indicator to each country-specific coronavirus shock. The figure shows that unemployment doesn’t necessarily need to skyrocket in response to health shocks. Unemployment insurance claims have reached record levels in the weeks after the outbreak in the US. The impulse responses are consistent with that knowledge, revealing an extremely large response of the Google unemployment indicator. Unemployment is also set to considerably increase in Spain. The country has a large share of workers on temporary contracts, who are more likely to become unemployed due to the uncertainty generated by the pandemic. Other countries in our sample, however, as well as the vast majority of countries not shown in the Figure, appear more successful in insulating their labor force from the crisis. Even if the recessionary effects on output are likely to be large everywhere, for most countries, early indicators of unemployment suggest that local labor markets are not going to experience the same turbulence as those in the U.S.

### 4.4 The Benefits of Social Distancing

So far, we have focused on the direction of causality that goes from COVID cases to social and economic responses. Here, we provide evidence on the opposite direction: the effects of social distancing on new COVID cases.

Figure 8 shows the impulse responses of the growth rate of COVID-19 cases in different countries to a social distancing shock, measured as a one-standard-deviation decline in social mobility. Social distancing leads to declines in the growth rate of coronavirus cases in the days after the shock. The only country in the Figure that doesn’t show a negative response is the U.K., for which social distancing has been much slower to start.

**Figure 8:**
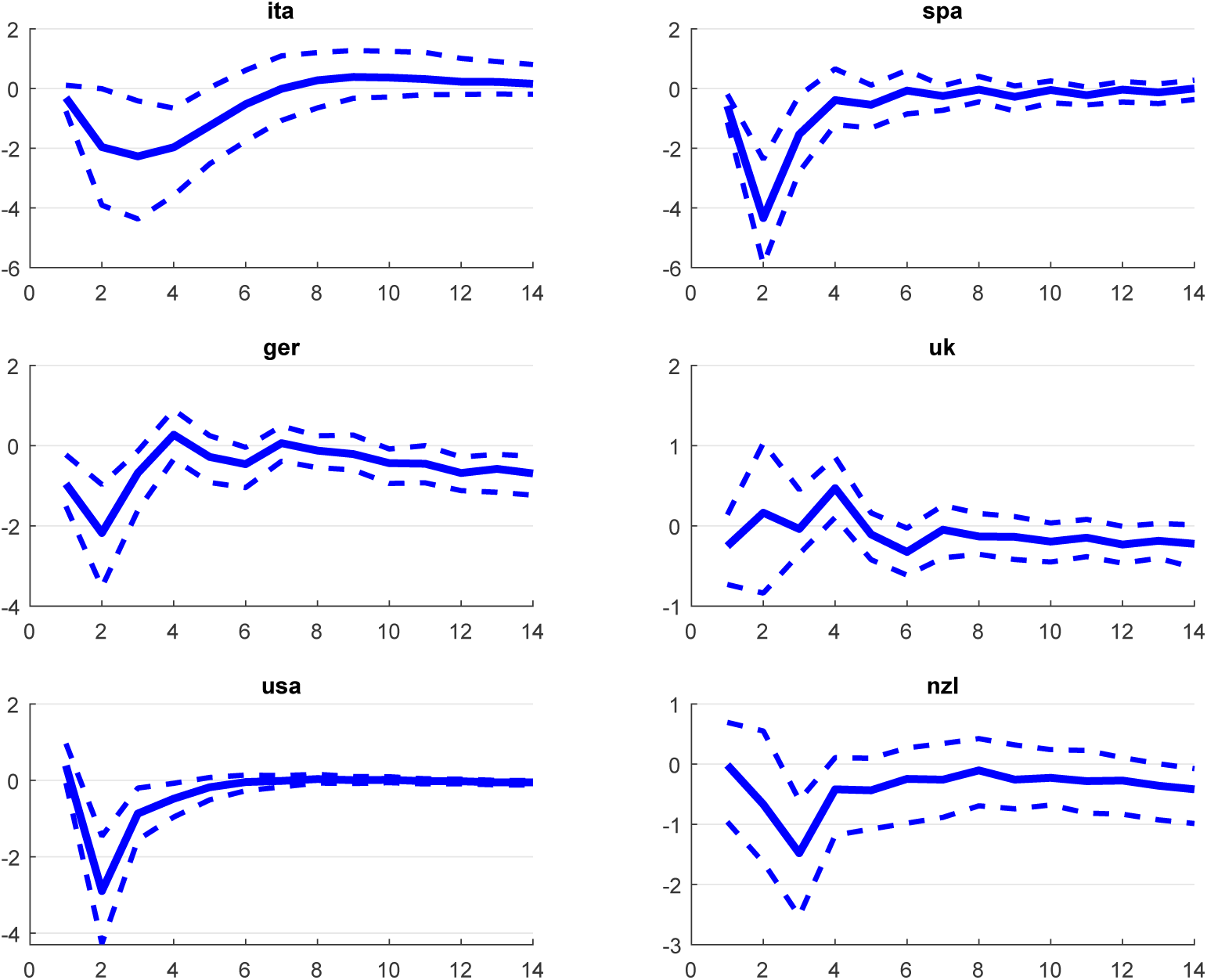
Impulse Responses of COVID-19 growth rates to a Social Distancing shock.

### 4.5 Discussion

Overall, this paper’s results highlight the importance of interconnections to understand not only the spread of the virus, but also adaptation and gradual learning in importing ideas and behavior from other countries. Risk perceptions and the willingness to engage in social distancing by the populations of most countries significantly respond to the corresponding variables in socially-connected countries. We stress the role of existing social networks across borders in the transmission of health shocks, perceptions about the risk of the disease, and ideas regarding the merit of social distancing.

The results reveal heterogeneous responses across countries to their own domestic coronavirus shocks. A common feature in all responses is that individuals responded with a lag and only gradually reduced their social mobility. This observation is consistent with epidemiological models that include adaptive human behavior and stress the role of informing and motivating people to reduce person-to-person contacts, such as the model presented in Fenichel et al. (2011).

Institutional differences among the countries’ labor markets are likely responsible for substantially different increases in unemployment. The lower degree of employee protections in the U.S. and the large share of temporary workers in the Spanish economy, are likely to account for the far worse outcomes in these countries. Everywhere else, fluctuations in unemployment have remained much tamer.

### 4.6 Sensitivity Analysis

In this section, we assess the sensitivity of our estimates to alternative data and econometric choices.

In the benchmark estimation, we have used data on COVID-19 cases transformed into daily growth rates. We can examine the sensitivity of the results to using the number of new daily cases, instead. We report in Table 2, the estimated interdependencies corresponding to those previously shown in Table 1. To save space, the results are shown for a subset of six countries. The estimates remain similar, with the exception of a smaller spillover of global risk into the domestic Italian risk perception variable.

**Table 2.** - Sensitivity Analysis: Contemporaneous effects of foreign aggregates. *Note*: sensitivity check i) repeats the estimation using the level of new daily COVID-19 cases, rather than their growth rate; case ii) estimates conditional vector-error-correction models rather than VAR for each country; case iii) computes changes in social mobility excluding the series related to residential mobility.

| Countries | COVID | Risk Perc. | Social Dist. | Countries | COVID | Risk Perc. | Social Dist. |
| --- | --- | --- | --- | --- | --- | --- | --- |
| i) <u>COVID-19 # of new cases instead of growth rate</u> |  |  |  |  |  |  |  |
| Italy |  | 0.559<br>(0.442) | 0.611***<br>(0.139) | U.S.A. |  | 1.617***<br>(0.157) | 0.545***<br>(0.117) |
| France | 3.818***<br>(1.080) | 0.792***<br>(0.152) | 0.519***<br>(0.102) | Germany | 5.242***<br>(0.996) | 0.785***<br>(0.194) | 1.655***<br>(0.175) |
| Spain |  | 1.677***<br>(0.232) | 1.103***<br>(0.222) | U.K. | 5.965***<br>(1.225) | 1.154***<br>(0.184) | 0.267*<br>(0.156) |
| ii) <u>VECMX* instead of VARX*</u> |  |  |  |  |  |  |  |
| Italy |  | 0.994**<br>(0.389) | 0.671***<br>(0.140) | U.S.A. |  | 1.619***<br>(0.150) | 0.527***<br>(0.108) |
| France | 1.052***<br>(0.131) | 0.863***<br>(0.157) | 0.702***<br>(0.095) | Germany | 1.041***<br>(0.108) | 1.111***<br>(0.207) | 1.703***<br>(0.189) |
| Spain |  | 1.528***<br>(0.234) | 1.200***<br>(0.241) | U.K. | 0.887***<br>(0.119) | 1.035***<br>(0.183) | 0.575***<br>(0.162) |
| iii) <u>Exclude Google Residential Mobility</u> |  |  |  |  |  |  |  |
| Italy |  | 1.061***<br>(0.384) | 0.652***<br>(0.154) | U.S.A. |  | 1.580***<br>(0.154) | 0.552***<br>(0.112) |
| France | 1.002***<br>(0.123) | 0.835***<br>(0.148) | 0.408***<br>(0.146) | Germany | 1.078***<br>(0.114) | 0.755***<br>(0.203) | 1.963***<br>(0.235) |
| Spain |  | 1.596***<br>(0.234) | 1.360***<br>(0.234) | U.K. | 0.924***<br>(0.121) | 1.085***<br>(0.185) | 0.500***<br>(0.151) |

Also, in our benchmark estimation, the conditional country-specific models corresponded to VARs with the addition of weighted foreign aggregates. Another option often used in the GVAR literature is to allow for long-run relationships and estimate Vector Error Correction (VECM) models instead.^6^ The results shown in Table 2, as well as all the main findings, remain in line with what discussed so far.

Finally, our Google mobility indicator was computed by taking the average of mobility changes across all available categories. It can be argued that the relevant social distancing measure that matters for health outcomes should exclude Residential mobility. Therefore, we repeat the analysis by constructing social mobility, but now excluding the residential component. Again, the results remain substantially unchanged.

## 5 Conclusions

We estimated a global model of 41 countries to examine the interconnections in coronavirus cases and social and economic responses over the first months of the COVID-19 pandemic.

The results suggest that social connections across borders are helpful in understanding the spread of the disease, but also the spread in perceptions and social behavior, among countries.

The outbreak led to risk perceptions about Coronavirus that differed internationally. The heterogeneous risk perceptions likely affected the countries’ social distancing responses. Populations in most countries, through their social networks, learned from experiences abroad and started to pay more attention to the evolving coronavirus situation and reduced their mobility. They displayed, however, a degree of behavioral adaption: they didn’t change their habits immediately, but only gradually over time. Some countries were outliers: they didn’t respond much through social distancing, to global or domestic shocks. As a result, they don’t show the same reduction in the growth rate of COVID-19 cases in response to social distancing that exists in other countries.

The original health shocks, either directly, or through increased uncertainty and social distancing, have economic effects. We focus, in particular, on the response of unemployment. While we do not have data at high frequency on the realized variable, we exploit daily data on an indicator that has been shown to predict actual unemployment quite accurately: unemployment from Google searches.

The response of unemployment across countries is very heterogeneous. In the US, unemployment skyrockets. This is consistent with the response of initial unemployment claims in the country. The same happens in Spain, with a large increase of unemployment in response to health shocks. In other countries with less precarious labor markets, the responses are more muted. While their economies are still likely to face a deep recession, fears of unemployment remain lower.

## Data Availability

All data are publicly available, with the exception of Facebook data, available through agreement with the company.

## Footnotes

1 For example, as documented in Brynildsrud and Eldholm (2020), the first cases in Nordic countries (in their case, Norway, but likely similar in neighboring countries) were due to travelers returning from vacations in Lombardy. To the extent that some of these tourism patterns increase the probability of Facebook links as well, which we believe reasonable, our measure will allow us to track likely routes for the spread of the disease.

2 The GVAR model has been proposed by Pesaran et al. (2004) and is surveyed in Chudik and Pesaran (2016).

3 The full list of countries is as follows: Australia, Austria, Belgium, Brazil, Canada, Chile, Colombia, Costa Rica, Czech Republic, Denmark, Estonia, Finland, France, Germany, Greece, Hungary, India, Indonesia, Ireland, Israel, Italy, Japan, Korea, Latvia, Lithuania, Luxembourg, Mexico, Netherlands, New Zealand, Norway, Poland, Portugal, Slovak Republic, Slovenia, South Africa, Spain, Sweden, Switzerland, Turkey, United Kingdom, United States. The only country that has been removed from the OECD list is Iceland, since Google mobility data were not available. For non-OECD key partners, we exclude China, since for our sample the numbers of cases had already declined.

4 Google LLC \Google COVID-19 Community Mobility Reports.” https://www.google.com/covid19/mobility/

5 For both Google Trends series, we use both the time series information and the cross-section information, by also extracting the relative popularity of the searches in each country for the period of interest. We then multiply the time series by the relative popularity in country *i* divided by the popularity in the country with the highest search volume (fixed at 100 in Google Trends by construction).

6 We preferred to refrain from cointegration relationships in our benchmark estimation, since it may be difficult to put confidence on long-run relationships estimated from two months of daily data.

## Notes

### Competing Interest Statement

The authors have declared no competing interest.

### Funding Statement

No funding.

